# A SARS-CoV-2 Delta Variant Containing Mutation in the Probe Binding Region Used for qRT-PCR Test in Japan Exhibited Atypical PCR Amplification and Might Induce False Negative Result

**DOI:** 10.1101/2021.11.15.21266335

**Authors:** Samiul Alam Rajib, Yasuhiro Ogi, Md. Belal Hossain, Terumasa Ikeda, Eiichi Tanaka, Tatsuya Kawaguchi, Yorifumi Satou

## Abstract

A recent pandemic of SARS-CoV-2 infection has caused severe health problems and substantially restricted social and economic activities. To cope with such an outbreak, the identification of infected individuals with high accuracy is vital. qRT-PCR plays a key role in the diagnosis of SARS-CoV-2 infection. The N protein-coding region is widely analyzed in qRT-PCR for the diagnosis of SARS-CoV-2 infection in Japan. We recently encountered two cases of SARS-CoV-2-positive specimens showing atypical amplification curves in the qRT-PCR. We performed whole-genome sequencing and found that the virus was a Delta-type variant of SARS-CoV-2 with a single nucleotide mutation in the probe-binding site. To evaluate the extent of spread of the variant in the area, we performed whole viral genome sequencing of samples collected from 61 patients infected with SARS-CoV-2 during the same time and in the same area. There were no other cases with the same mutation, indicating that the variant had not spread in the area. Furthermore, we performed phylogenetic analysis with various SARS-CoV-2 sequences deposited in the public database. Hundreds of variants were reported globally, and one in Japan were found to contain the same mutation. Phylogenetic analysis showed that the variant was very close to other Delta variants endemic in Japan but quite far from the variants containing the same mutation reported from outside Japan, suggesting that the variant would have been sporadically generated in some domestic areas. These findings propose two key points: i) mutations in the region used for SARS-CoV-2 qRT-PCR can cause abnormal amplification curves; therefore, the qRT-PCR result should not just be judged in an automated manner, but also manually checked by the examiner to prevent false-negative results, and ii) various mutations can be generated sporadically and unpredictably; therefore, efficient and robust screening systems are needed to promptly monitor the emergence of *de novo* variants.

## INTRODUCTION

Since its first report from a seafood market in Wuhan, China, in 2019 [1,2], coronavirus disease 2019 (COVID-19) has spread over six continents within a short period; the World Health Organization (WHO) declared COVID-19 a global pandemic in March 2020 [3]. As of November 4, 2021, there have been more than 247.4 million confirmed cases of COVID-19 worldwide, with over 5 million deaths attributed to it [4]. A metagenomic RNA sequencing study by Wu et al. in 2020 revealed that the virus causing COVID-19 is a novel strain of coronavirus, closely associated with severe acute respiratory syndrome coronavirus (SARS-CoV) and the middle east respiratory syndrome coronavirus (MERS-CoV). Therefore, it was named “severe acute respiratory syndrome coronavirus 2 (SARS-CoV-2)” [2].

The whole viral genome sequence of SARS-CoV-2 was first reported in January 2020 [2]. Since then, several quick and reliable diagnostic tests have been developed, including quantitative reverse transcription-polymerase chain reaction (qRT-PCR)-based diagnostic methods [5,6], CRISPR/Cas12 based methods [7], specific high-sensitivity enzymatic reporter unlocking (SHERLOCK) assay [8], and loop-mediated isothermal amplification (LAMP)[9]. To date, the United States Food and Drug Administration (USFDA) has issued emergency use authorization (EUA) for 266 molecular diagnostic tests that were developed based on the sequence information provided by the SARS-CoV-2 genome [10]. Although each method has advantages and disadvantages, qRT-PCR based methods have been the most common diagnostic methods applied to detect SARS-CoV-2 in patient samples. Several protein-coding sequences of the SARS-CoV-2 genome have been used for qRT-PCR, such as genes encoding RNA-dependent RNA polymerase (RdRP) [11], Spike (S), Nucleocapsid (N) [12] and Envelope (E) proteins [13].

In Japan, the multiplex Quantitative reverse-transcription PCR (qRT-PCR) based diagnostic method was first developed by the National Institute of Infectious Disease (NIID) in 2020 [14] and applied for the mass diagnosis of SARS-CoV-2 in 2020 [15]. NIID suggested the N1 and N2 regions of the N protein-coding sequences as the target for qRT-PCR amplification (Figure 1A). Since then, 26.6 million tests have been performed in Japan, among which 6% were positive for COVID-19 [16]. When the study was conducted, 124,358 diagnostic tests were performed in Kumamoto Prefecture in Japan, of which 14,392 tests were positive [17].

**Figure 1:**
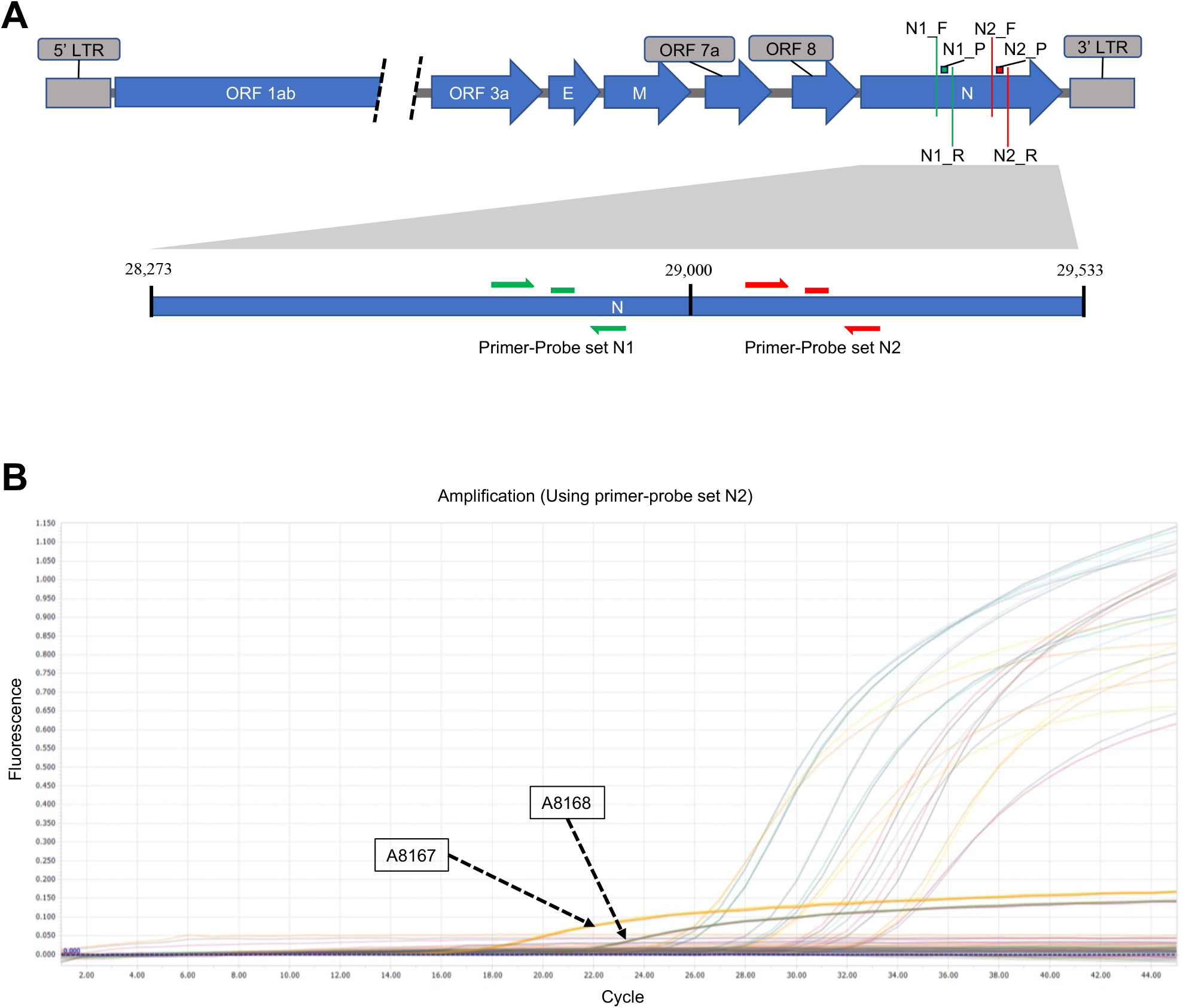
**(A)** Location of SARS-CoV-2 genome where the qRT-PCR primer-probe sets target for detecting SARS-CoV-2 in patient samples. Both primer-probe sets (N1 and N2) target the N protein-coding sequences. N1_F, N1_R, N1_P are the primers and probe of the N1 set and N2_F, N2_R. N2_P are the primers and probe of the N2 set (Figure not drawn to scale). **(B)** qRT-PCR curve to detect SARS-CoV-2 using NIID recommended N2 primer-probe set produced aberrant curve in two samples (A8167 and A8168) compared to other positive samples in the same run.

qRT-PCR based testing plays a vital role in identifying infected patients and confining them where uninfected individuals can perform social activities [18]. Therefore, accuracy in detecting SARS-CoV-2 in patients plays a crucial role in controlling the transmission of this highly contagious pathogen. False-negative results put much stress on the containment efforts for COVID-19. Successful qRT-PCR based detection is dependent on the efficient binding of primers and probes to the target areas. Any changes in the target nucleotide sequence can significantly lower the binding affinity of the primers and probes to their target region, resulting in a pseudo-negative diagnosis [19,20]. Consequently, the resulting undetected mutants can also result in future waves of COVID-19 [21]. Although SARS-CoV-2 has a lower mutation frequency compared to other RNA viruses because of its RNA proofreading mechanism [22], its higher transmission rate makes SARS-CoV-2 a good candidate to observe for any unforeseen mutation within the qRT-PCR primer and probe binding regions so that the quality of the detection methods could be ensured.

This article reports a point mutation in the *N* gene of SARS-CoV-2 that resulted in an atypical qRT-PCR curve during the diagnostic test, leading to a dubious diagnostic interpretation. Sequencing results revealed the presence of a single point mutation at the probe-target site in the *N* gene. This analysis shows that sequencing can play a problem-solving role in qRT-PCR based diagnostic complications and raise caution to institutes performing qRT-PCR tests to detect SARS-CoV-2 infection.

## MATERIALS AND METHODS

### Regulatory Approvals

In this study, samples were collected by the Kumamoto City Medical Association Inspection Center (hereinafter referred to as the Kumamoto PCR center). The downstream analyses were performed at the Division of Genomics and Transcriptomics in the Joint Research Center for Human Retrovirus Infection, Kumamoto University [Approved by the Ethics Review Committee of the Faculty of Life Sciences Kumamoto University (Approval no. 2223)].

### Specimen collection and storage

For this study, sputum and/or nasopharyngeal swab samples were collected from the Kumamoto city area from August 16, 2021, to September 8, 2021, as part of the regular COVID-19 diagnostic service. The collected samples and extracted RNAs were preserved at -80 °C until further experiments were performed according to the guidelines of the NIID, Japan [23].

### RNA extraction and qRT-PCR

For qRT-PCR analysis and sequencing, RNA was extracted from the sputum and/or nasopharyngeal swab samples (140 µL) using the QIAamp Viral RNA Mini Kit (QIAgen) and EZ1 Virus Mini Kit v2.0 (QIAgen) according to the manufacturer’s instructions. The final RNA product was eluted in 60 µL of buffer AVE (RNase free water with 0.04% sodium azide), and 5 µL of the eluted RNA was used for qRT-PCR, and the remaining RNA was stored at -80 °C for further analysis. qRT-PCR was performed on a LightCycler96 System (Roche, Basel, Switzerland) according to the protocol suggested by Shirato et al. [14] to detect the *N* gene using the One Step PrimeScript^™^ RT-PCR Kit (Perfect Real Time) (Takara). The N2 primer-probe set was used as described by Shirato et al. [14].

### SARS-CoV-2 whole-genome sequencing

cDNA synthesis, viral sequence enrichment, library amplification, and indexing were performed using the QIAseq DIRECT SARC-CoV-2 kit (QIAgen), following the manufacturer’s recommendations. For multiplexing the samples, QIAseq DIRECT UDI set-A (QIAgen) was used. SARS-CoV-2 libraries of 25 µL volume for each sample were prepared at the end of the process. The quality of the enriched SARS-CoV-2 libraries was evaluated by electrophoresis with a TapeStation 4150 system (Agilent Technologies). Finally, the prepared libraries were denatured and subjected to sequencing using MiSeq reagent Micro and Nano Kits (Version 2, 300 cycles) in the MiSeq desktop sequencing system (Illumina).

### Data analysis

Upon sequencing in the Illumina MiSeq sequencer, one FASTQ file (Read1) was generated for each sample. Adapter sequences were trimmed from the Read1 using the ‘cutadapt’ tool [24]. After the adapter trimming step, a cleaning step was performed using the PRINSEQ tool [25] and Read1 with Phred score > 20 were used for downstream analysis. Adapter trimmed and cleaned reads were then aligned to the SARS-CoV-2 reference genome NC_045512 (isolate Wuhan-Hu-1) using the BWA-MEM algorithm [26]. Subsequently, the Samtools program [27] was used to remove multiple aligned reads, and Freebayes (Version 1.2.0) command-line tools [28] were used to call the variants from the aligned reads and create a variant call format (VCF) file. Finally, the aligned files and VCF files were visualized using the Integrative Genomics Browser (IGV) [29]. The resulting consensus SARS-CoV-2 genomes were deposited at the Global Initiative on Sharing All Influenza Data (GISAID) [30] (Supplementary data 1).

For secondary data analysis, the Pangolin COVID-19 Lineage Assigner tool was used to assign the nomenclature for the viral genomes proposed by Rambaut et al. [31] and the CoV-GLUE web application tool was used to check for any novel mutations within the viral sequences [32].

### Phylogenetic tree analysis

For local phylogenetic tree construction, 61 SARS-CoV-2 strains were collected from Kumamoto city between September 1, 2021, and September 8, 2021. First, the viral sequences were aligned to the SARS-CoV-2 reference genome NC_045512 (isolate Wuhan-Hu-1) using Geneious Prime (version 2020.2.4) (https://www.geneious.com/). Then, the aligned sequences were used to construct a maximum-likelihood phylogenetic tree using phyML 3.0 [33] with Smart Model Selection (SMS) [34].

SARS-CoV-2 genomes and associated metadata were downloaded from the GISAID database [30] (accessed on October 18 2021) to construct a global phylogenetic tree. Global genome collections were downloaded from the “Region-specific Auspice source file” of the GISAID database, resulting in 3580 viral genomes (globally distributed random viral strains collected from December 2019 to October 2021) for global phylogenetic tree construction. Globally distributed viruses with the G29234A mutation were identified using the ‘substitution’ tool from the GISAID database (accessed on October 18 2021) [30]. Then, the resulting 250 viral genomes containing the G29234A mutation were added to the Nextstrain build [37] as ‘focal’ sequences. The “global genome collections” were kept as ‘contextual’ genomes to build the phylogenetic tree. No non-human viral hosts were considered for tree construction.

## RESULTS

### Real-time PCR showed an atypical curve

NIID initially recommended a multiplex qRT-PCR system for detecting SARS-CoV-2 using two primer and probe sets (N1 and N2) [14]. Subsequently, according to the 3^rd^ edition of “Guidelines for the operation of the new coronavirus (SARS-CoV-2) test methods, NIID recommends using N2 primer-probe set with one enforcement to detect viral RNA in the samples during the epidemic period in which outbreaks continue in multiple municipalities [38]. Following the instructions from NIID, the Kumamoto PCR center used the N2 primer-probe set for diagnosis purposes. In this qRT-PCR experiment, 20 samples collected from the Kumamoto city area in the same time frame (Supplementary data1) were run in duplicate. From the qRT-PCR curve, it was seen that two of the samples had lower C_*t*_ values (Figure 1, Table 1); however, the fluorescence intensity of samples A8167 and A8168 was not amplified in the same way as the other samples during the same run (Figure 1). This unusual amplification of the qRT-PCR curve gave rise to the possibility of some variations in the primer-probe target sequences of the *N* gene of those samples.

**Table 1:** Cycle threshold (C_*t*_) values of 20 samples collected from Kumamoto city area from August 16, 2021, to September 7, 2021.

| Sample | $C_t$ values | |
| --- | --- | --- |
|  | Read 1 | Read 2 |
| A5631 | 33.78 | 33.83 |
| A5715 | 33.19 | 33.15 |
| A5806 | 35.14 | 34.65 |
| A5993 | 32.92 | 32.92 |
| A6131 | 30.11 | 30.28 |
| A6203 | 28.35 | 28.32 |
| A6338 | 33.39 | 33.46 |
| A6450 | 29.05 | 29.02 |
| A6563 | 32.23 | 32.5 |
| A6844 | 30.58 | 30.57 |
| A6881 | 27.12 | 27.03 |
| A6954 | 30.13 | 30.06 |
| A6979 | 28.86 | 28.81 |
| A7122 | 27.27 | 27.25 |
| A7123 | 31.15 | 31.11 |
| A7410 | 26.26 | 26.26 |
| A7456 | 32.04 | 31.92 |
| A7885 | 30.38 | 30.4 |
| A8167 | 20.19 | 20.02 |
| A8168 | 24.3 | 24.36 |

### SARS-CoV-2 whole-genome sequencing revealed G29234A mutation

Based on the hypothesis mentioned earlier, whole-genome sequencing was performed initially on seven samples (five samples showing characteristic qRT-PCR curve and two samples showing aberrant qRT-PCR curve from the same run) following the protocol mentioned in the Materials and Methods section. IGV visualization of the VCF files revealed the presence of a G→A mutation at 29234 loci of the SARS-CoV-2 genome of A8167 and A8168 samples, whereas no mutation was found in the same locus for the rest of the samples (Figure 2A). Coverage depth analysis revealed that the reads covering 29,234 loci were enriched with mutated base A (n=2296, 99.9%), whereas the wild type base G was supported by only three reads (0.1%) for sample A8167 (Figure 2B). A similar pattern was observed in sample A8168 as well, where the mutated A base was supported by 1,066 reads (100%) compared with the wild type G base with zero (0%) reads in the mentioned position of the SARS-CoV-2 genome (Figure 2B). This mutation overlaps with the *N* gene probe used in the recommended N2 primer-probe set (Figure 1A). It can be assumed that the G29234A mutation in samples might be a cause of the aberrance in the qRT-PCR curve by considering the sequencing data obtained from the samples and their corresponding qRT-PCR curve.

**Figure 2:**
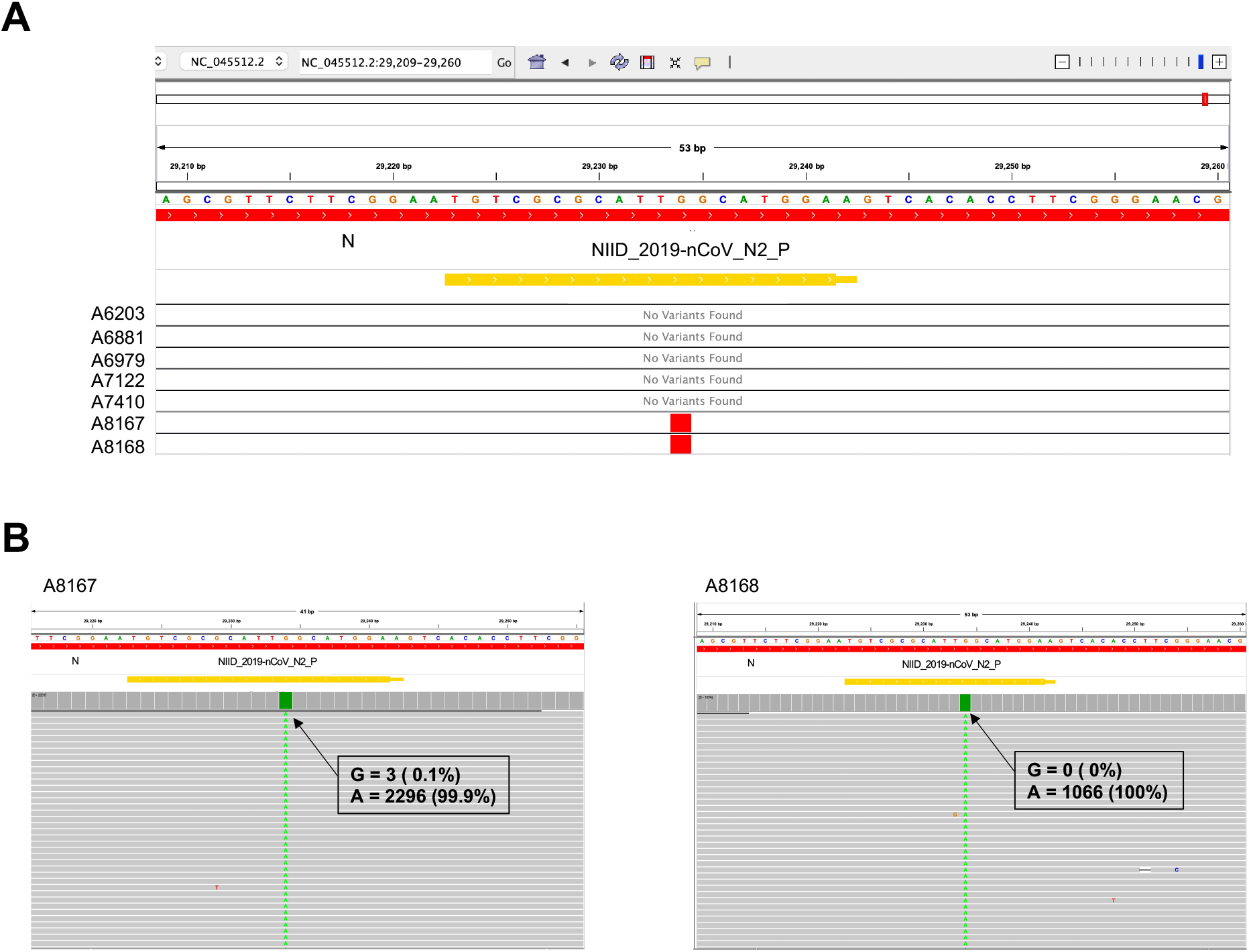
**(A)** Integrative Genomics Viewer (IGV; https://software.broadinstitute.org/software/igv/) screenshot depicts the Variant Call Format (VCF) file showing point mutation for the seven sequenced samples. Single nucleotide mutations were observed at the 29234 loci of the SARS-CoV-2 genome for both A8167 and A8168 samples. The orange bar labelled “NIID_2019-nCOV_N2_P” corresponds to the region covered by the N2 probe suggested by Shirato et al. [14]. **(B)** The BAM files show the reads and coverage for A8167 (left) and A8168 (right). Numbers in the box correspond to the raw read number and the percentage of the reads at the 29234 loci of the SARS-CoV-2 genome.

### Phylogenetic analysis showed the similarities among the local strains in Kumamoto

Whole-genome sequencing of another 61 samples collected from Kumamoto city within the same time frame was performed to investigate whether the variant containing the G29234A mutation was endemic or sporadic in the Kumamoto area. The resulting sequences showed no variation in the 29234 loci of the SARS-CoV-2 genome (Supplementary Figure 1). For further confirmation, a local phylogenetic tree was constructed using the variants containing the G29234A mutation and the variants without the mutation collected from Kumamoto city within the same time frame (Figure 3). The variants containing the G29234A mutation clustered within the same clade (AY29; Delta variant Japanese sub-lineage [39]) in the local phylogenetic tree. The horizontal branch distances in the tree did not deviate much for the mutated samples when compared with the rest of the samples from the same clade. These results confirmed that the mutant samples (A8167 and A8168) are genetically similar to the local strains but differ in mutation landscape and that the G29234A mutation was sporadic at the time of reporting in the local area.

**Figure 3:**
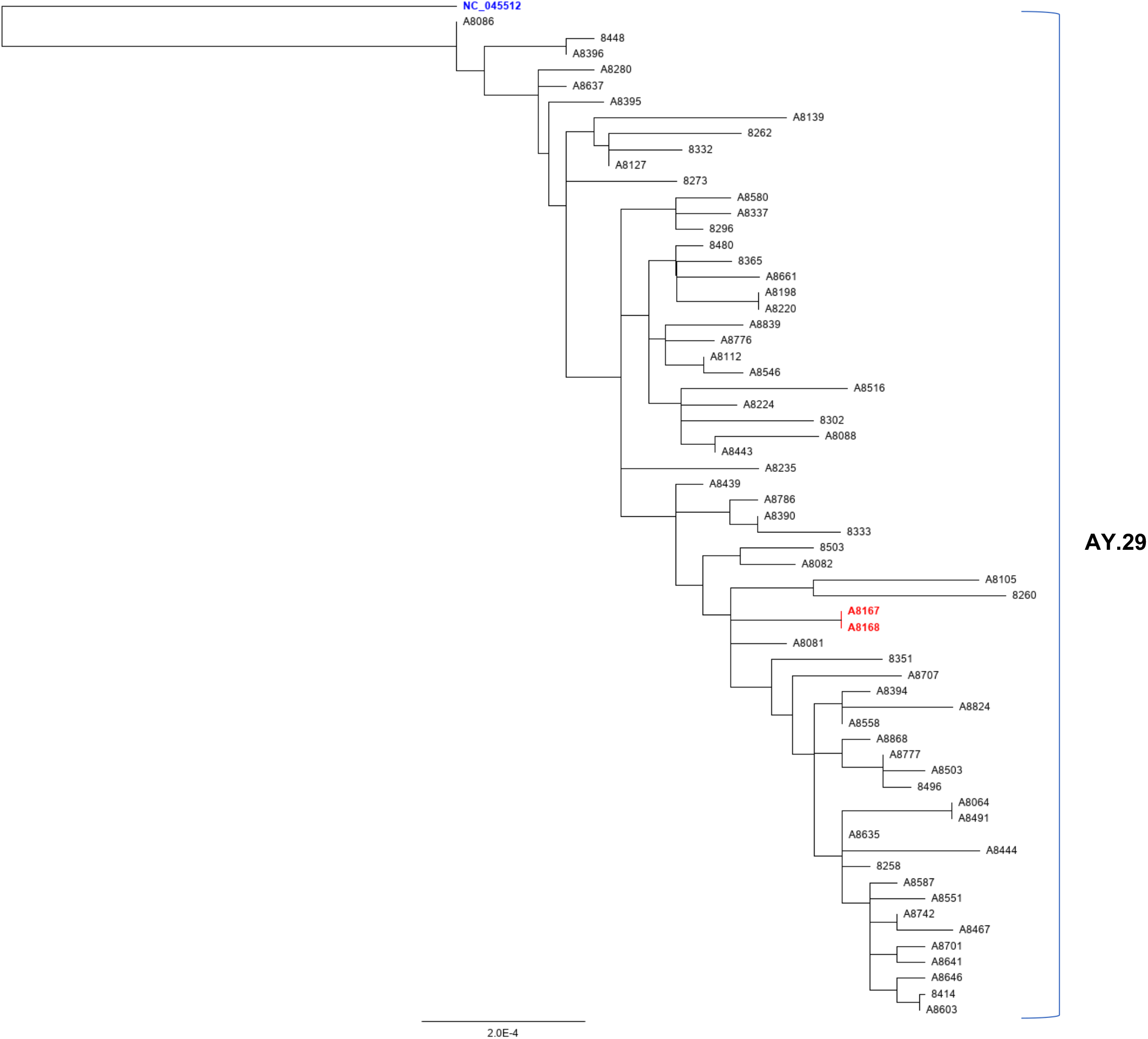
Maximum likelihood phylogenetic tree constructed using Geneious prime (version 2020.2.4) (https://www.geneious.com/prime/) with 63 locally collected samples from Kumamoto city. The samples represent the same locality withing the same time frame. The scale bar indicates the number of substitutions per site. The Wuhan-hu-1 (NC_045512; heighted in blue) is placed as the root sequence of the tree and the red-highlighted samples contain the G→A mutation at 29234 loci of SARS-CoV-2 genome. AY is the alias used for B.1.617.2 lineage (Delta variant)

Next, SARS-CoV-2 sequences deposited in the GISAID database were analyzed to determine whether the same mutated variant was distributed across different geographical locations. When highly covered genomes (> 29,000 bp), excluding the low-coverage genomes (> 5% Ns), were searched for the mutations, 250 samples were found to have the same G→A mutation at 29234 loci from 3,341,006 submitted genomes to the GISAID database (mutation frequency 0.007%; accessed on October 18, 2021). After creating the global phylogenetic tree with randomly sampled sequences from different locations across the world along with the mutated viral samples, several endemic clusters were identified throughout the course of the SARS-CoV-2 pandemic (Figure 4). Initially, a sporadic variant group with G29234A mutation was observed in Canada in April 2020, followed by two isolated clusters in England and Germany between December 2020 and June 2021. Nevertheless, the same mutation was observed in groups mainly concentrated in the USA and Europe in recent times (Figure 4). Another phylogenetic tree was constructed with the globally distributed G29234A variants and the variants sampled in the Kumamoto city containing the G29234A mutation (Figure 5) to understand a possible source of the mutated variant found in Kumamoto city. The resultant tree clearly showed cluster separation between the G29234A variants sampled from the rest of the world and the samples collected from the Kumamoto city (Figure 5; zoomed in-right panel). Considering the factors such as relative cluster position of G29234A mutants collected from the Kumamoto area (Figure 3, Figure 5) to the other mutant strains in the tree and the sample collection dates (Supplementary data2), it could be inferred that the mutated variants might not have migrated from outside. Instead, they acquired these mutations sporadically in Japan.

**Figure 4:**
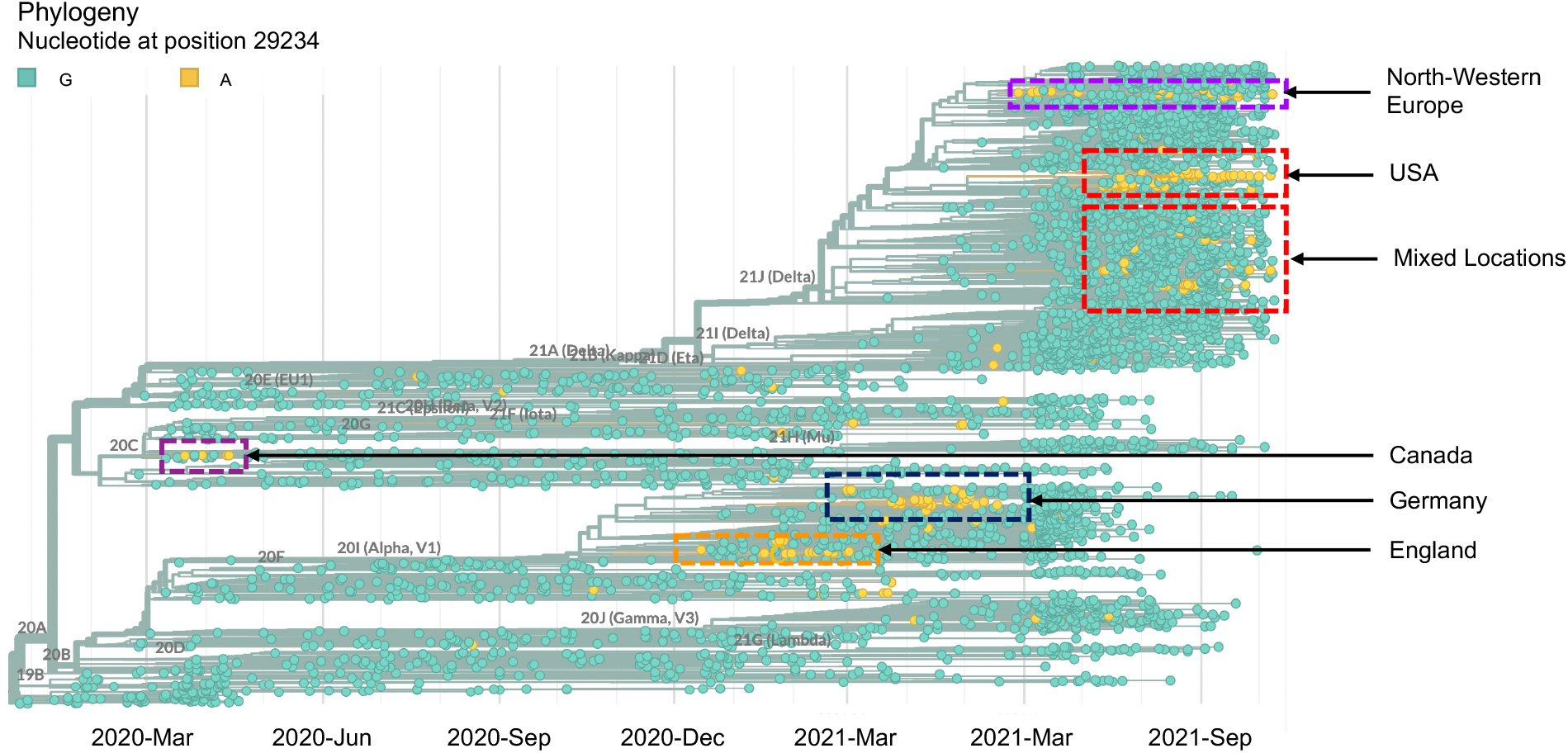
Nextstrain [37] generated phylogenetic tree with viral strains containing A allele at 29234 loci of the SARS-CoV-2 genome (highlighted in orange).

**Figure 5:**
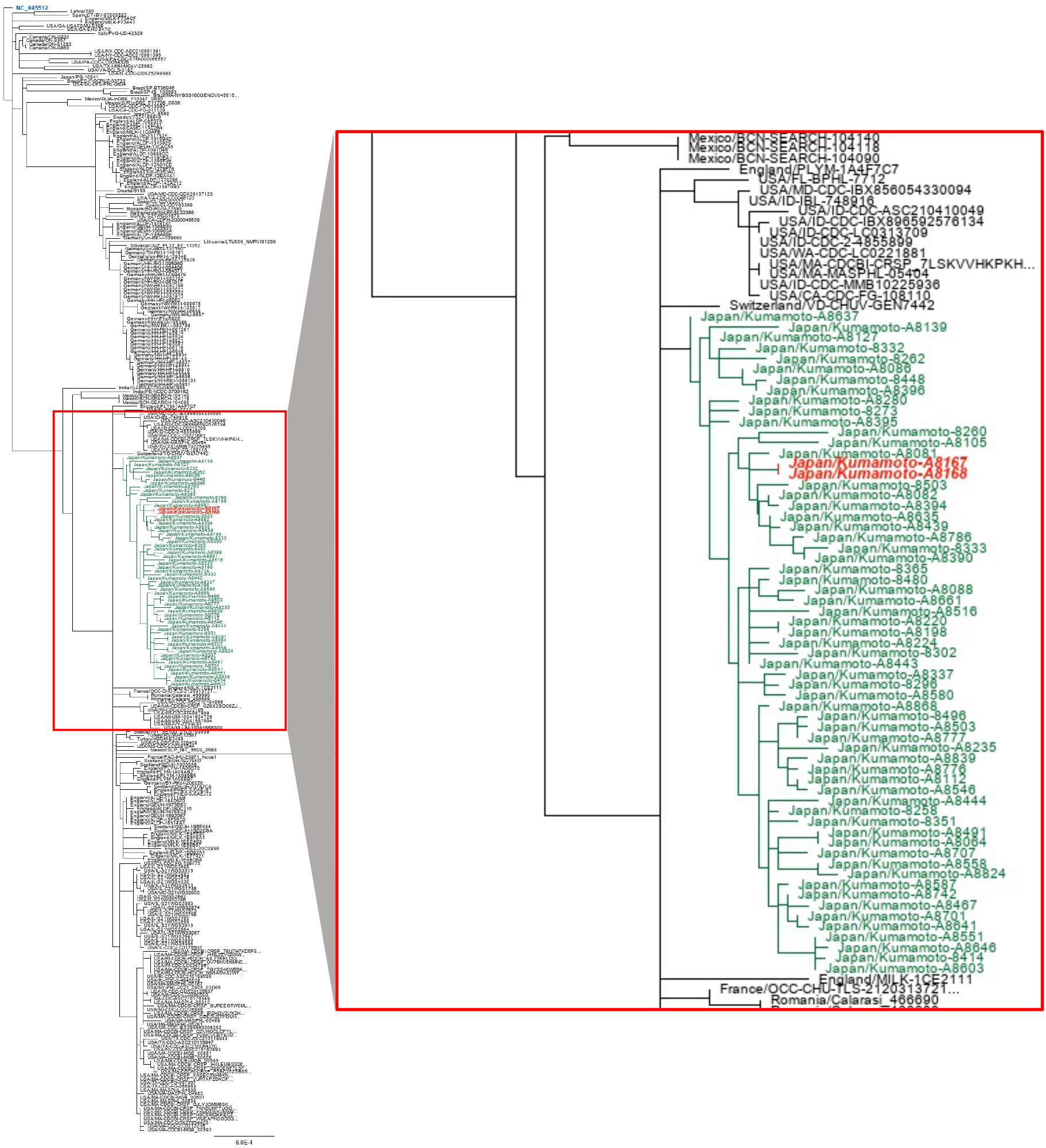
Maximum likelihood phylogenetic tree built in Geneious prime (version 2020.2.4) (https://www.geneious.com/prime/) showed the relationships of viral strains collected from Kumamoto area from August 16, 2021 to September 8, 2021 with the viral strains having G29234A mutation from around the world. The Wuhan-hu-1 (NC_045512; heighted in blue) is placed as the root sequence of the tree and the green-highlighted samples represented the viral strains collected from the Kumamoto city. Red-bold faced viral strains (Zoomed in-Right panel) represent viral strains collected from Kumamoto area which contain G29234A mutation.

## DISCUSSION

From the position of primers and probes suggested by Shirato et al. [14], it could be said that the presence of G→A mutation at 29234 bp [position relative to reference sequence NC_045512.2] played a role in obtaining the abnormal qRT-PCR curves while diagnosing the patients using the NIID recommended N2 primer-probe set in Kumamoto PCR center. According to the “Guidelines for the operation of the new coronavirus (SARS-CoV-2) test methods (3^rd^ Edition)”, the sensitivity of the N2 primer-probe set was higher than that of the N1 set with all test samples during the protocol development. Similar results were found even in samples with low viral titers [38]. Therefore, the NIID advised all test centers to follow their recommended operation instructions during different endemic conditions. NIID recommended using the N2 primer-probe set with one enforcement during the epidemic period in which outbreaks continue in multiple municipalities [38]. Therefore, the similar use of the N2 primer-probe set might have been adopted in other prefectural test centers during the ongoing epidemic waves in Japan. Given this situation, the possibility of the presence of the G29234A mutation in the circulating strains might jeopardize some test results by returning a dubious qRT-PCR curve [19,20].

Initially, the G29234A mutation showed a homoplasy trait. It appeared spontaneously in two clades at two time points: in 20C (clustered in Canada) during the early pandemic period in 2020 and 20I (also known as Alpha strain) from December 2020 to June 2021, mainly concentrated in England and Germany (Figure 4). However, from the recent trait, it was seen that the mutation had been spreading among the Delta variants of SARS-CoV-2. Although the frequency of this mutation was 0.007% compared to the total submitted genomes in the GISAID database at the time of reporting, the existence of this mutation in multiple clusters among the 21J (Delta) clade (Figure 4, Supplementary Figure 2) in different geological locations at the same time raised the possibility of local spreading. It also increased the probability that the mutations observed in the Kumamoto area might have migrated from external sources. However, the phylogenetic analysis revealed that the strains that acquired the G29234A mutation in Japan were distantly located in a separate cluster with other strains collected from the Kumamoto area compared to those with the same mutation in other regions of the world (Figure 5), which suggested sporadic acquisition of the mutations in Japan instead of being migrated from other external sources.

While searching for a possible causal mechanism for the G292324A mutation, we found a 5′-UGG (edited G is underlined) trinucleotide motif in the genomic strand of SARS-CoV-2. This complementary strand is 5′-CCA (edited C is underlined), suggesting the sequence preference of RNA for the APOBEC3G cytosine deaminase [40,41]. However, at least three studies with SARS-CoV-2 genomic sequence analyses have shown that changes in C→U are biased in the viral genomic stand [42–44]. Therefore, the G292324A mutation may not be due to deamination by APOBEC3 proteins. Further investigation is required to confirm the involvement of APOBEC3 enzymes and/or other mechanisms in the G292324A mutation. In addition, the G29234A mutation caused a change in the amino sequence from glycine (G) to serine (S) at 321-position of the N protein. To determine why this mutation in the N protein did not sustain the other clades during the early pandemic and why this mutation had been spreading within the Delta variants requires further investigation.

The above findings indicate two key points. First, mutations in the primer-probe target regions could cause an atypical qRT-PCT curve, leading to false-negative results. Second, various mutations can occur sporadically and unpredictably. Therefore, efficient and robust screening systems are deemed necessary to promptly monitor the emergence of new variants of interest.

## Acknowledgements

We are grateful to Y. Matsuoka and E. Harada for technical support and valuable discussions; A. Iemura and T. Kashiwagi for their dedicated efforts to establish and run the PCR Center, Kumamoto City Medical Association Inspection Center. We also thank Dr Yuki Furuse from the Institute of Frontier and Life Sciences, Kyoto University, for his valuable comments on the analysis and ‘Editage’ for English language editing.

This work was supported by research grants from the Japan Agency for Medical Research and Development (AMED) (JP21fk0410023, JP21wm0325015, JP21jm0210074) to Y. Satou, the JSPS Leading Initiative for Excellent Young Researchers (LEADER), Japan Science and Technology Agency (JST) A-STEP (JPMJTM20SL), Shin-Nihon Foundation of Advanced Medical Research and an intramural grant from the Kumamoto University COVID-19 Research Projects (AMABIE) to T. Ikeda. The funders had no role in the study design, data collection, data interpretation, or discussion regarding submission for publication.

## Author approval

The authors confirm that the manuscript has been read and approved by all named authors and that there are no other persons who satisfied the criteria for authorship but are not listed. The authors further confirm that the order of authors listed in the manuscript has been approved by all.

## Data availability statement

The SARS-CoV-2 genome sequences were downloaded from GISAID, which are subject to GISAID’s terms and conditions (https://www.gisaid.org/registration/terms-of-use/).

---

**Supplementary Figure 1:**
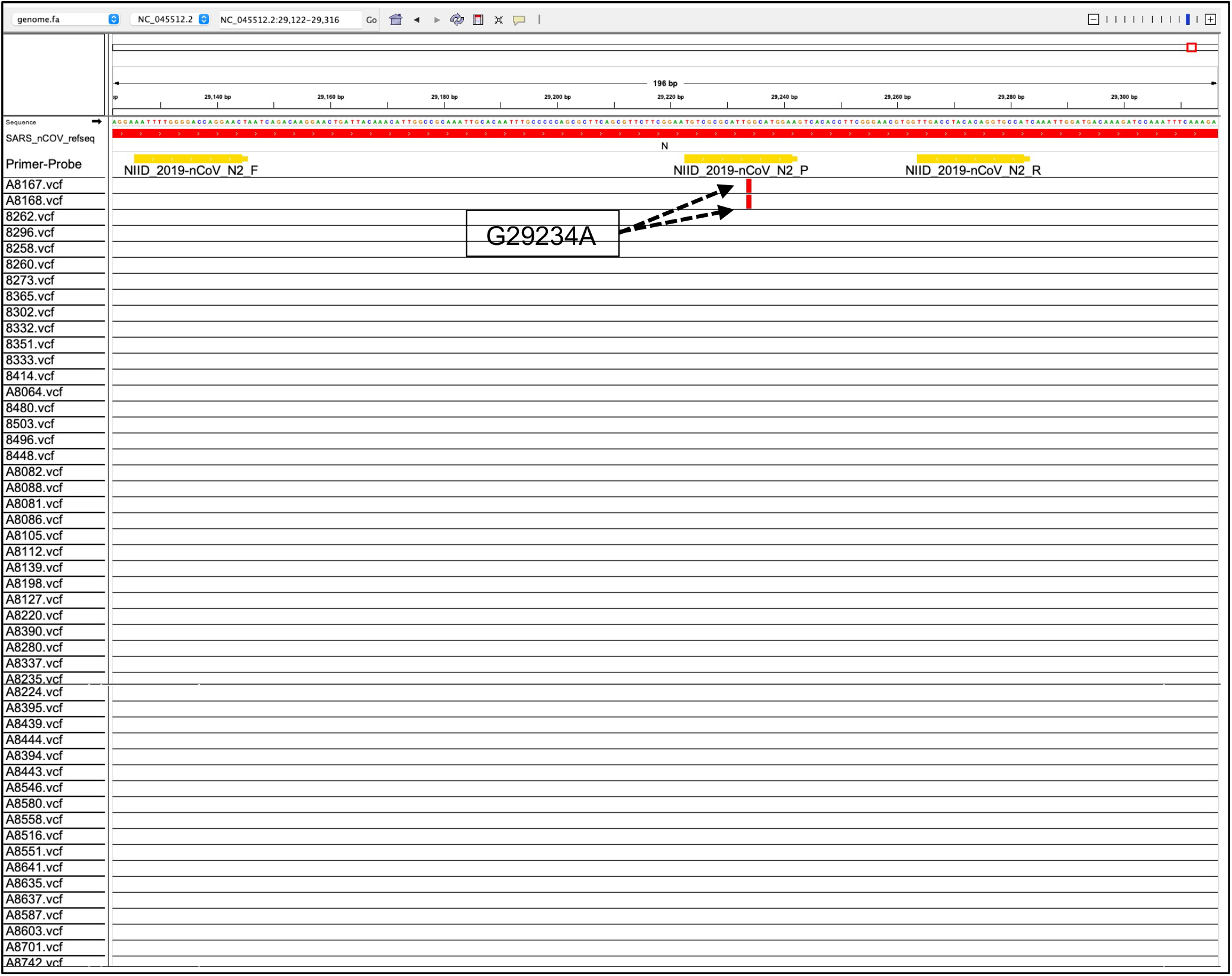
Visualization of Variant Call Format (VCF) files of 63 samples (2 mutated samples and 61 other samples) collected from Kumamoto city area from September 1, 2021, to September 8, 2021, using Integrative Genomic Viewer (IGV). The red bar corresponding to the sample name depicts the presence of a mutation in that particular sample and the particular position corresponding to the reference sequence (NC_045512.2). The orange bar labelled “NIID_2019-nCOV_N2_P” reaches the region covered by the N2 probe suggested by Shirato et al. [14].

**Supplementary Figure 2:**
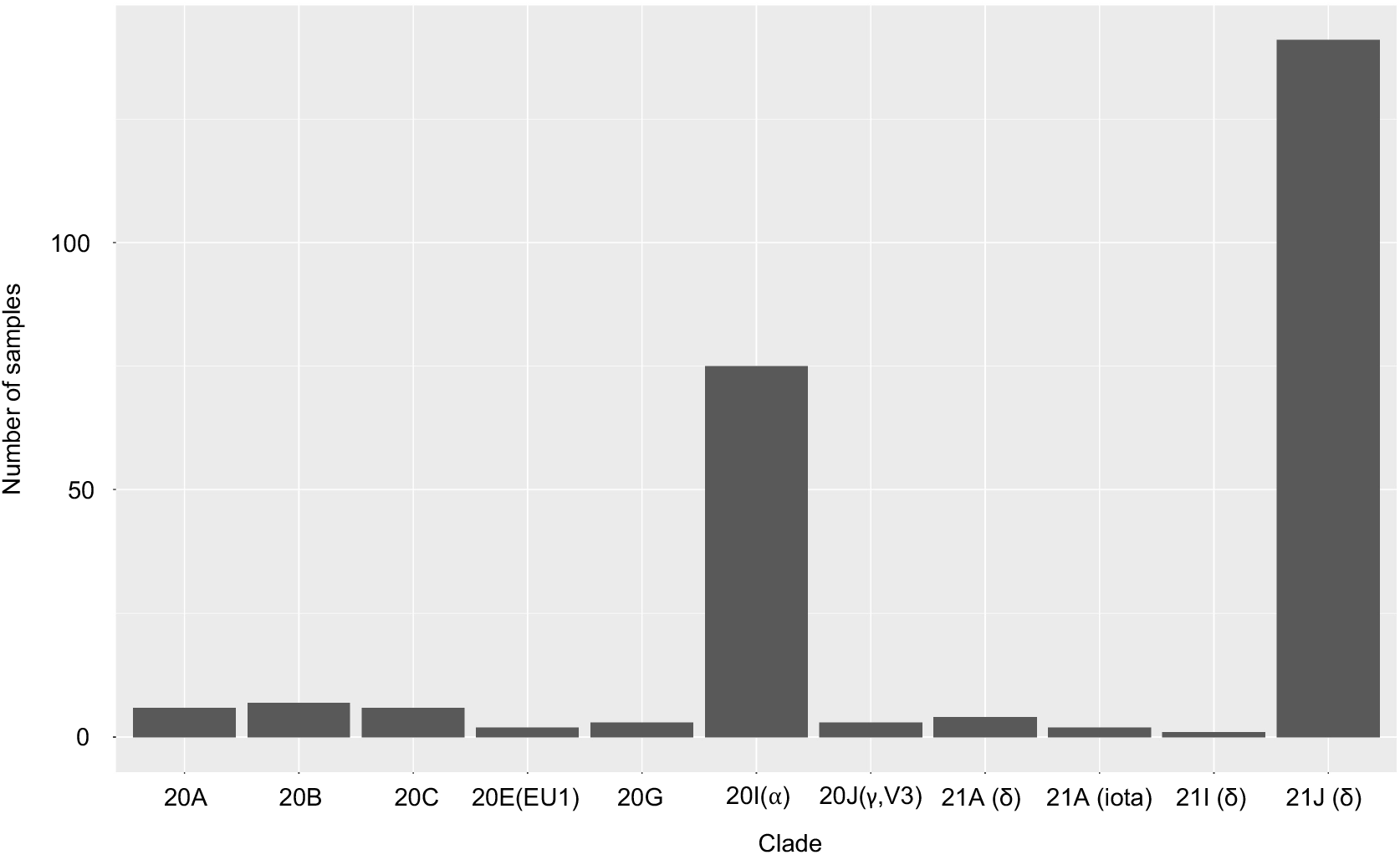
Distribution of G29234A mutants among the clades. Clades are defined according to Nextstrain clade naming and definitions (https://docs.nextstrain.org/projects/ncov/en/latest/reference/naming_clades.html). Variants of concern (VOC) are marked according to the Greek alphanumeric symbols within the bracket adjacent to the relevant Nextstrain clade names.

